# Cost-effectiveness analysis comparing repetitive transcranial magnetic stimulation therapy with antidepressant treatment in patients with treatment-resistant depression in Japan

**DOI:** 10.1101/2022.12.02.22282995

**Authors:** Yoshihiro Noda, Chiaki Miyashita, Yoko Komatsu, Shinsuke Kito, Masaru Mimura

## Abstract

**Background:** In Japan, repetitive transcranial magnetic stimulation (rTMS) for patients with treatment-resistant depression (TRD) became covered by the National Health Insurance (NHI) in 2019. While the usefulness of rTMS for TRD has been established, the cost-effectiveness of rTMS versus antidepressants has not been thoroughly analyzed in Japan. This study aimed to evaluate the cost-effectiveness of rTMS for TRD under the NHI system.

**Methods:** Cost-effectiveness of rTMS versus antidepressants was analyzed using a microsimulation model to compare the direct costs and quality-adjusted life years (QALYs) in patients with moderate to severe depression who had failed one or more antidepressants over their lifetime. Model inputs of clinical parameters and the utility were derived from published literature. Cost parameters were estimated from the Japanese Claim Database. In addition, the robustness of the analysis results was evaluated using sensitivity analysis and scenario analysis.

**Results:** The analysis estimated that rTMS increased effectiveness by 0.101QALYs and total cost by ¥94,370 ($689) compared with antidepressant medications. As a result, the incremental cost-effectiveness ratio (ICER) of rTMS was estimated to be ¥935,984 ($6,832)/QALY. In the sensitivity and scenario analyses, ICER did not exceed ¥5 million ($36,496)/QALY as the reference value of the Japanese public cost-effectiveness evaluation system.

**Limitations:** For the portion of the rTMS for which data were not available under the NHI system in Japan, foreign data and estimates were extrapolated.

**Conclusions:** rTMS showed the potential to be a cost-effective treatment strategy for TRD compared with antidepressants under the NHI system in Japan.

## Introduction

Depression is a disorder deeply related to the reduction of the patient’s quality of life (QOL) (Lépine and Briley, 2011). According to the Global Burden of Disease Study 2017 (GBD 2017 Disease and Injury Incidence and Prevalence Collaborators, 2018), depression was ranked among the three leading illnesses in terms of the years lost due to disability (YLDs) associated with the disorder. In addition, depression causes a loss of productivity, resulting in significant social losses and a negative impact on the economy (Asami et al., 2015; Yamabe et al., 2019). The global population with depression is estimated to be 280 million (3.8%) (World Health Organization, 2021), and the number of patients with affective disorders including depression in Japan is estimated to increase to 1.72 million in 2020 (Ministry of Health, Labour and Welfare of Japan, 2020), according to a patient survey by the Ministry of Health, Labor and Welfare.

Approximately 30% or more of patients with depression do not respond to appropriate antidepressant medications, and such cases are termed treatment-resistant depression (TRD) (Furukawa et al., 2000; Rush et al., 2006). The annual medical cost for patients with TRD is over $6,000 larger than that of patients with treatment-responsive depression, and the frequency of hospitalization is approximately twice as high as for treatment-response depression (Mrazek et al., 2014). Thus, not only the mental and physical burdens but also the financial burden is greater for patients with TRD.

Most of guidelines recommend pharmacotherapy, psychotherapy, pharmacotherapy in combination with psychotherapy, and electroconvulsive therapy as acute phase of treatments (Gelenberg et al., 2010; Lefaucheur et al., 2014; Milev et al., 2016; Perera et al., 2016). Especially for patients with moderate to severe depression, antidepressant treatment is recommended. In addition, recently, repetitive transcranial magnetic stimulation (rTMS) therapy is also recommended as one of the first to second-line alternatives to pharmacotherapy an effective, non-invasive, and safe treatment for TRD, supported by a high level of evidence (George et al., 2010). In Japan, rTMS therapy for adult patient with moderate or severe TRD was approved by the Japanese regulatory authority in 2017 and was listed as a therapy reimbursed by the National Health Insurance (NHI) in 2019.

Hitherto, cost-effectiveness analyses of rTMS therapy in patients with TRD in the United States and Australia showed that compared with pharmacotherapy (Nguyen and Gordon, 2015; Voigt et al., 2017), rTMS therapy is highly effective and even reduces medical costs. As such, rTMS has also been reported to have medical-economic benefits in several countries (Nguyen and Gordon, 2015; Voigt et al., 2017). Though studies reporting the efficacy of rTMS for TRD have gradually appeared in Japan (Ikawa et al., 2022), no study has been published that quantitatively analyzed and evaluated rTMS from a medical economic perspective. Currently, in Japan, rTMS is reimbursed only for use in acute phase treatment of the same episode with the price of ¥12,000 per session ($88US: $1US = ¥137 as of August 25, 2022 (International Monetary Fund, 2022)). In addition, the NHI system does not cover the use of rTMS for continuous treatment for patients who have had insufficient response to acute phase treatment, or for maintenance phase treatment to prevent recurrence. Therefore, the application of rTMS in Japan significantly differs from that in other countries, which makes it difficult to simply compare the economic benefits of rTMS in other countries with those in Japan. TRD is currently a national health problem and the socioeconomic impact of TRD is enormous in Japan. Thus, quantitative evaluation of the health economic benefits of rTMS under the NHI system provides important information for determining the selection and provision of appropriate health care services for patients with TRD. Hence, the objective of this study was to evaluate the cost-effectiveness of rTMS therapy compared with antidepressants as an acute phase treatment for patients with TRD under the framework of NHI-covered healthcare in Japan.

## Methods

### Overview

The study used a microsimulation model to estimate lifetime medical costs and quality-adjusted life years (QALYs) for patients with moderate or severe TRD who did not respond to initial antidepressant medication in the first treatment step. Estimates for this analysis were made under the following two strategies: (1) switch to rTMS therapy as an acute phase treatment in the second treatment step and (2) continue receiving antidepressant medication from the first treatment step. This was followed by calculation of the incremental cost-effectiveness ratio (ICER) for rTMS therapy in comparison with antidepressant medication.

### Model structure

The microsimulation model was constructed using TreeAge Pro 2022, R2.0 (TreeAge Software, Williamstown, MA, USA). Patients entered the model and transitioned to the various health states which were defined as acute phase treatment, maintenance phase treatment, and termination of treatment based on their responsiveness to their therapies as response, remission, relapse, and recurrence. The period from acute phase treatment to remission or termination of treatment was defined as a single treatment period (Figure 1). Since maintenance phase treatment with rTMS is not yet reimbursed by the NHI, rTMS was only applied as the acute phase treatment in step 2 and switched to pharmacotherapy as maintenance phase treatment after the patient responds. The comparator was a case in which pharmacotherapy was conducted during the acute phase of step 2 treatment and continued for the maintenance phase. During each step, treatment was continued if the patient showed temporary remission in response to acute phase treatment, and treatment was terminated if no worsening of symptoms was observed for a certain period. For patients who responded to acute phase treatment but did not show remission, antidepressant medication was continued for another month before proceeding to the next treatment step. Treatment was switched to step 3 antidepressant medication when patients have failed to respond to rTMS therapy administered as step 2 of acute phase treatment or showed partial response to the same therapy but did not achieve remission. In addition, transition to step 3 was considered when patients showed temporary remission but developed exacerbation of symptoms before the termination of treatment, and they achieved remission with treatment but developed recurrence after the termination of treatment. After step 3, the model was set up with no difference in treatment selection between patients who received rTMS and patients who received antidepressant medication in the acute phase of treatment of step 2. A subset of patients who proceeded to step 5 were set up to receive hospitalized treatment and management. After hospitalization in step 5, patients who did not respond to treatment, who responded to treatment but did not achieve remission, or who developed symptom exacerbation after remission, were set to receive electroconvulsive therapy (ECT). Patients who achieved remission with inpatient treatment and ECT in step 5 were to be treated and managed on an outpatient basis unless their symptoms exacerbated. For patients who did not respond to ECT and for patients who partially responded to ECT but did not achieve remission, life-long cycles of 6 months of inpatient treatment & management and 6 months of outpatient treatment & management was set up. Patients who did not receive hospitalized care in step 5 were to continue to be treated and managed in an outpatient setting. Transition to death was considered for each health state. A total of 2 million patients were simulated in the base case analysis using a microsimulation.

**Figure 1.**
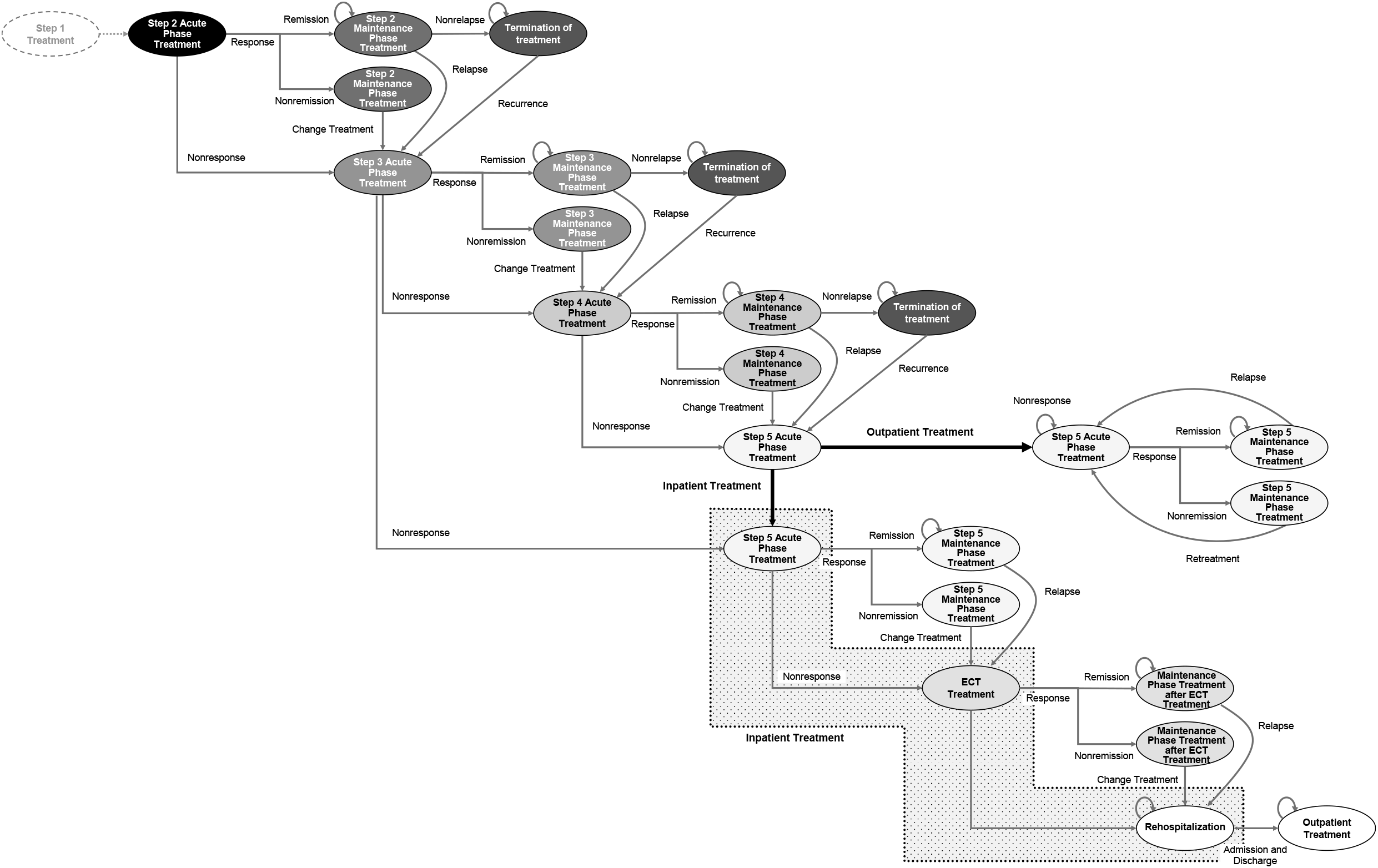
Model Structure. The microsimulation model was constructed. Patients entered the model and transitioned to the various health states which were defined as acute phase treatment, maintenance phase treatment, and termination of treatment based on their responsiveness to their therapies as response, remission, relapse, and recurrence. The period from acute phase treatment to remission or termination of treatment was defined as a single treatment period. Transition to state of death was considered for each health state.

### Parameters

#### Clinical parameters

Clinical parameters identified from the published literature are listed (Table 1). Response and remission rates for rTMS therapy were derived from the NeuroStar® Advanced Therapy System Clinical Outcomes Registry Study, which evaluated the efficacy of rTMS therapy for patients with TRD in clinical settings across the United States (Sackeim et al., 2020). Recurrence rates were derived from the randomized controlled trial comparing efficacy in three groups (rTMS plus antidepressants, rTMS therapy alone, and antidepressants alone) in patients with moderate to severe depression who had shown remission or partial remission following 6 months of pharmacotherapy (Wang et al., 2017). Here, since no previous studies explicitly reported the recurrence rate of rTMS therapy for depression, we assumed that the relapse and recurrence rates were equivalent in this study. On the other hand, response, remission, and recurrence rates for each treatment step in antidepressant therapy were obtained from the STAR*D study, which assessed the efficacy of each treatment line in patients with nonpsychotic depression receiving switching antidepressant administration (Rush et al., 2006). The STAR*D study reported rates of response, remission, and recurrence up to step 4, but not on each parameter after step 5. Therefore, the value of each parameter in step 5 was regarded as equivalent to the parameters in step 4. Recurrence rates for each treatment step in antidepressant therapy were based on the results of the study by Hardeveld et al. that investigated recurrence rates in patients with depression in the Netherlands (Hardeveld et al., 2013) as well as the studies that compared cost-effectiveness among antidepressants in patients with TRD (Wang et al., 2017; Young et al., 2017). Clinical parameters of ECT were cited from the cost-effectiveness analysis of ECT compared with antidepressants for patients with depression (Ross et al., 2018). The percentage of patients who commenced inpatient treatment when there was no response in steps 3 or 4 and the percentage of patients who commenced inpatient treatment in step 5 were estimated from the Japanese Claims Database analysis. Regarding the mortality rate of patients with depression, we assumed that the overall mortality rate of patients with depression would not be different from that of the general population, although the number of deaths due to suicide is relatively high in depression. Based on this assumption, data from the Abridged Life Tables 2021 (Ministry of Health, Labour and Welfare of Japan, 2021a) were used for mortality rates of patients with depression. The duration of maintenance phase treatment from the remission to the termination of treatment was set based on the expert opinions of the three authors (Y.N., S.K., and M.M.) who are specialists in the field in light of the current situation in clinical psychiatry in Japan.

**Table 1.**
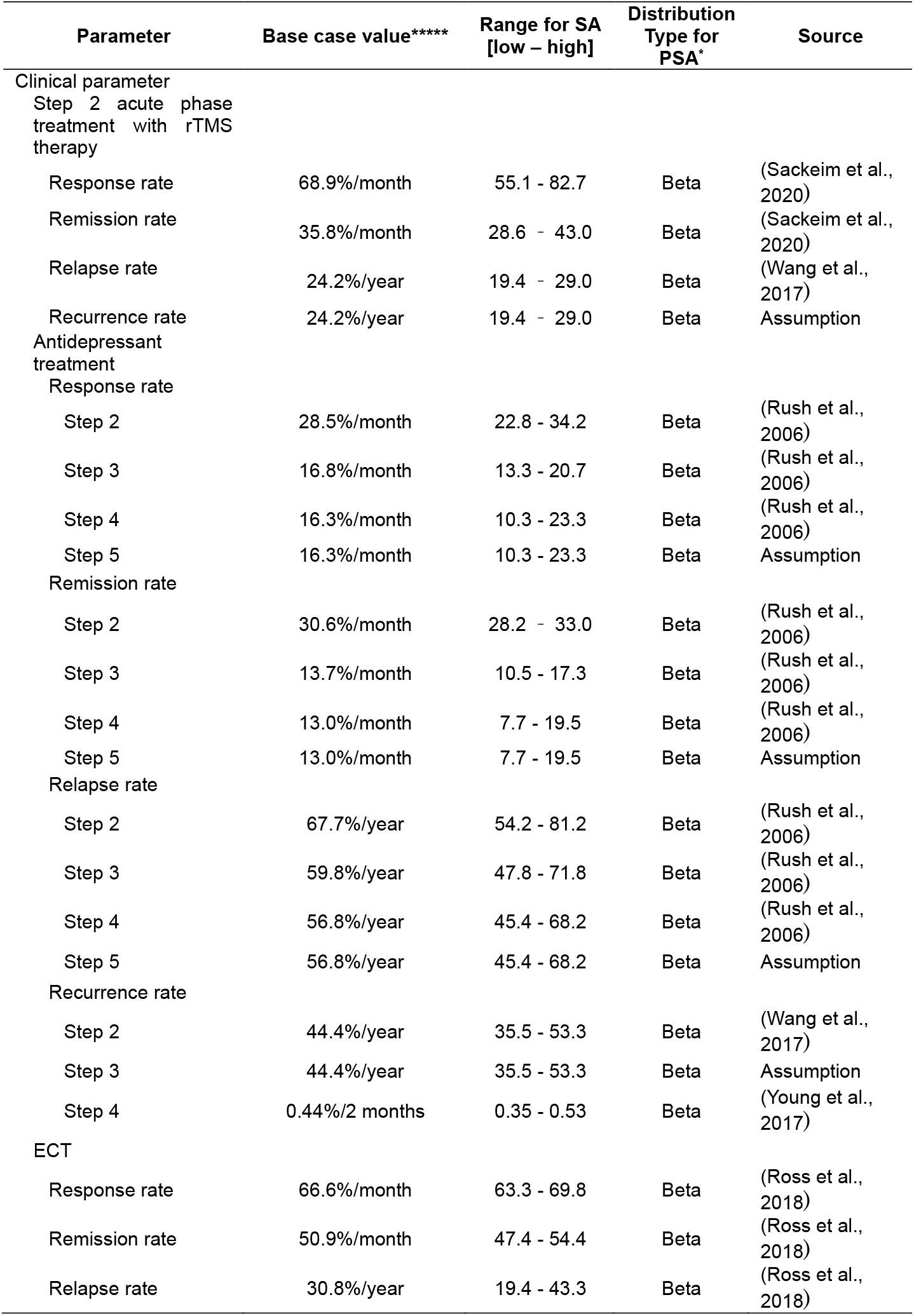
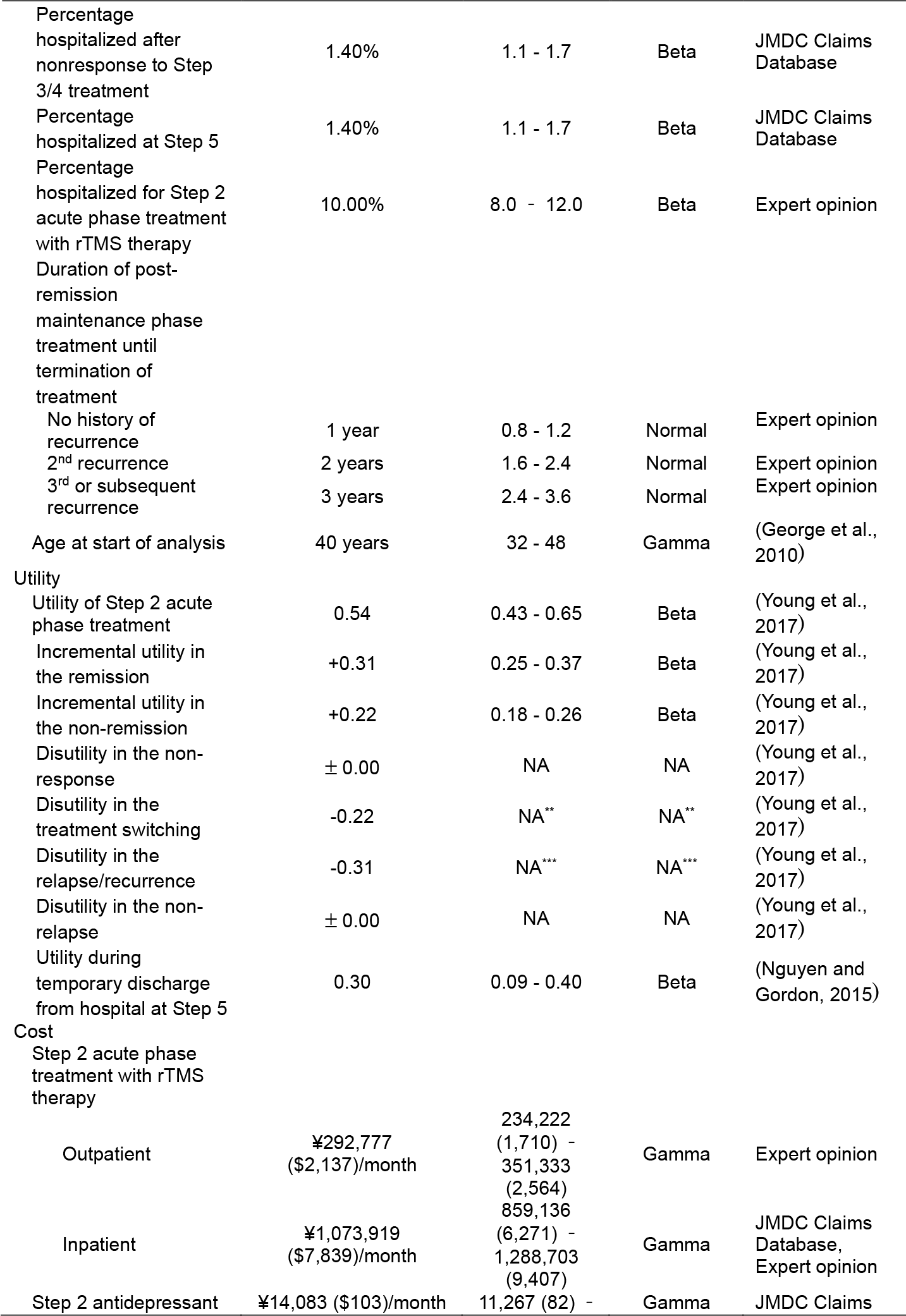
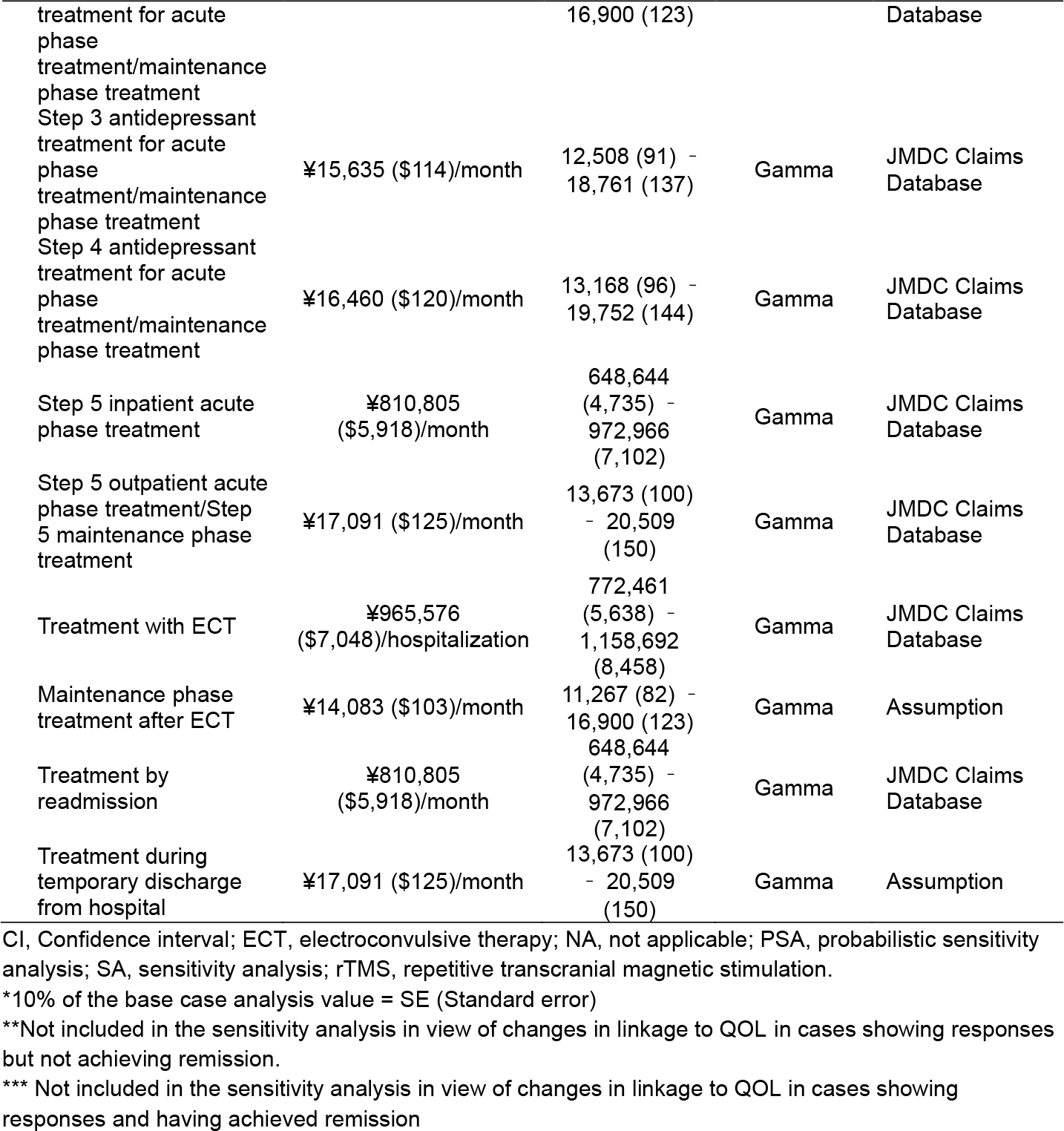
List of Parameters.

#### Utility parameters

Patient utility values were identified from the published literature (Table 1). The utility values for patients with depression were derived from the values estimated in the study by Young et al. (Young et al., 2017), and the utility values after admission in patients were derived from the previous study on the cost-effectiveness of rTMS (Nguyen and Gordon, 2015).

#### Resources / treatments costs

The direct costs of each treatment were based on the JMDC Claims Database (JMDC Inc., Tokyo, Japan). This claims database consists of inpatient and outpatient claims data provided by insurers (health insurance societies) (Table 1). The database includes healthcare information for a cumulative observation population of approximately 7.3 million people, as of February 2020, from more than 90 health insurance societies. We used the claims data from patients diagnosed with a depressive episode (ICD-10 code: F32) or repetitive depressive disorder (ICD-10 code: F33) from July 2016 to June 2019.

The definition of TRD for the database analysis was based on patients who were eligible for rTMS therapy (Supplementary Figure 1). The database analysis included analysis of the costs for antidepressant treatment, inpatient treatment for depression, outpatient examination, and hospitalization to receive ECT for each patient with TRD at each treatment step (Supplementary Table 1.,2,3). The cost for rTMS therapy was estimated from insurance reimbursement prices, assuming five weekly treatments. rTMS therapy does not require hospitalization, but patients receiving this therapy can be hospitalized in part because of the need for medical management or simply because of accessibility issues to rTMS therapy. There was insufficient data on the percentage of patients receiving rTMS therapy on an inpatient basis. However, given the fact that rTMS therapy is generally administered in outpatient settings overseas and based on the expert opinions of the authors, we set step 2 acute phase treatment with rTMS therapy in an inpatient setting at 10% of the total number of patients.

### Analytical Methods

We conducted the microsimulation as the starting age for the analysis was 40 years old. Simulation analysis was performed over the lifetime of each patient in one-month cycles. The analysis was conducted from the perspective of public healthcare payers, and only direct medical costs were considered. The effectiveness was evaluated using QALYs, while the cost-effectiveness was evaluated using ICER. Both the costs and effectiveness were discounted at 2% per year.

### Scenario analysis

The following five scenario analyses were conducted: 1) patients who remitted after the acute phase treatment of rTMS in step 2 are transferred to weekly outpatient treatment of rTMS as maintenance phase until either transition to step 3 or the termination of treatment; 2) patients who have a recurrence after completion of the acute phase treatment in step 2 receive reintroduction of rTMS therapy as the acute phase treatment in step 3; 3) patients receive rTMS plus antidepressant treatment in acute phase of step 2; 4) the loss of productivity due to depression in patients is considered; 5) the time horizon is changed to 10 years (Supplementary Table 4).

### Sensitivity analysis

A one-way sensitivity analysis was conducted to confirm the influence of each parameter on the analysis results, with the results represented by a tornado diagram. For the range of variation of parameters, the discount rate was set to 0% to 4%, and for other parameters, in the absence of statistical information on 95% confidence intervals, the base case value ±20% was applied. A probabilistic sensitivity analysis with 1,000 Monte Carlo simulations was performed to assess the uncertainty of the analytical results. For the probability distribution of each parameter, a gamma distribution was assigned to the cost parameters and a beta distribution was assigned to the probability parameters and utilities (Briggs et al., 2006). When statistical information on the variance of each parameter was not available, we used a theoretical distribution with a standard error of 10% of the base case value.

## Results

The effectiveness for patients with TRD was 21.627 QALYs when rTMS therapy was applied as step 2 acute phase treatment, and 21.526 QALYs when antidepressant treatment was continued. The incremental effectiveness of rTMS therapy was 0.101 QALYs compared with antidepressant treatment. The cost during the step 2 acute phase treatment was ¥428,558 ($3,128) with rTMS therapy and ¥46,398 ($339) with antidepressant treatment. Thus, the step 2 acute phase treatment costs increased by $382,161 ($2,789) due to rTMS therapy. When rTMS therapy was applied as step 2 acute phase treatment, the number of patients who required hospitalization decreased, resulting in a decrease in hospitalization costs of ¥118,192 ($863). In terms of total cost over the lifetime, rTMS therapy as step 2 of acute phase treatment costs ¥9,040,065 ($65,986) while continued antidepressant treatment as the same step costs ¥8,945,695 ($65,297), indicating that rTMS therapy as step 2 of acute phase treatment increased total costs by ¥94,370 ($689). As a result, the ICER with rTMS therapy in comparison with antidepressant treatment for step 2 acute phase treatment was ¥935,984 ($6,832)/QALY (Table 2).

**Table 2.** Results of Base Case Analysis.

| Variable | rTMS therapy | Antidepressant treatment | Incremental |
| --- | --- | --- | --- |
| Total cost/patient, yen (\$) | 9,040,065 (65,986) | 8,945,695 (65,297) | 94,370 (689) |
| Step 2 (Acute phase treatment & maintenance phase treatment)/patient, yen (\$) | 428,558 (3,128) | 46,398 (339) | 382,161 (2,789) |
| Step 3 (Acute phase treatment & maintenance phase treatment)/patient, yen (\$) | 35,621 (260) | 34,613 (253) | 1,008 (7) |
| Step 4 (Acute phase treatment & maintenance phase treatment)/patient, yen (\$) | 37,127 (271) | 36,032 (263) | 1,095 (8) |
| Step 5 (Outpatient care)/patient, yen (\$) | 20,272 (148) | 20,745 (151) | -473 (-3) |
| Step 5 (Inpatient care)/patient, yen (\$) | 5,261,196 (38,403) | 5,431,713 (39,648) | -170,517 (-1,245) |
| ECT treatment/patient, yen (\$) | 28,437 (208) | 29,150 (213) | -713 (-5) |
| After readmission/patient, yen (\$) | 3,228,853 (23,568) | 3,347,044 (24,431) | -118,192 (-863) |
| QALYs per patient | 21.627 | 21.526 | 0.101 |
| ICER, yen (\$)/QALY | - | - | 935,984 (6,832) |

ECT, electroconvulsive therapy; ICER, incremental cost-effectiveness ratio; QALYs, quality-adjusted life years; rTMS, repetitive transcranial magnetic stimulation

Table 3 summarizes the results of scenario analyses. Three scenarios showed that QALYs gained with rTMS were higher than those with antidepressants, but the costs were less. One was that reintroduction of rTMS therapy at the time of recurrence after completion of step 2 acute phase treatment. The second one was concurrent use of rTMS and antidepressant treatment as step 2 acute phase treatment. The third one was consideration of productivity loss due to depression. The other scenario analyses showed the ICER was ¥2,418,868 ($17,656)/QALY for the scenario where rTMS therapy was continued as step 2 maintenance phase treatment while the ICER was ¥1,513,265 ($11,046)/QALY for the scenario in which the time horizon was changed to 10 years.

**Table 3.**
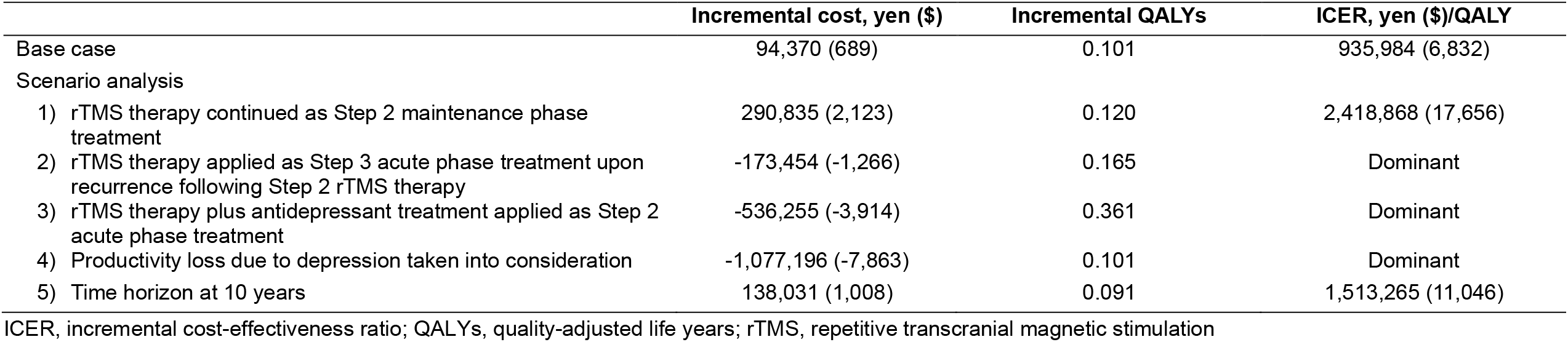
Results of Scenario Analysis.

| | Incremental cost, yen (\$) | Incremental QALYs | ICER, yen (\$)/QALY |
| --- | --- | --- | --- |
| Base case | 94,370 (689) | 0.101 | 935,984 (6,832) |
| Scenario analysis |  |  |  |
| 1) rTMS therapy continued as Step 2 maintenance phase treatment | 290,835 (2,123) | 0.120 | 2,418,868 (17,656) |
| 2) rTMS therapy applied as Step 3 acute phase treatment upon recurrence following Step 2 rTMS therapy | -173,454 (-1,266) | 0.165 | Dominant |
| 3) rTMS therapy plus antidepressant treatment applied as Step 2 acute phase treatment | -536,255 (-3,914) | 0.361 | Dominant |
| 4) Productivity loss due to depression taken into consideration | -1,077,196 (-7,863) | 0.101 | Dominant |
| 5) Time horizon at 10 years | 138,031 (1,008) | 0.091 | 1,513,265 (11,046) |

ICER, incremental cost-effectiveness ratio; QALYs, quality-adjusted life years; rTMS, repetitive transcranial magnetic stimulation

Of the parameters set as the model for the one-way sensitivity analysis, the one that had the greatest impact on the analysis results was the remission rate of step 2 acute phase treatment with rTMS therapy (Figure 2). Furthermore, given the ICER of ¥5 million ($36,496)/QALY which is the reference value for the public cost-effectiveness evaluation system in Japan (Center for Outcomes Research and Economic Evaluation for Health, 2019), the results of the probabilistic sensitivity analysis indicated that the probability of rTMS therapy for TRD to be cost-effective was 100% (Figure 3).

**Figure 2.**
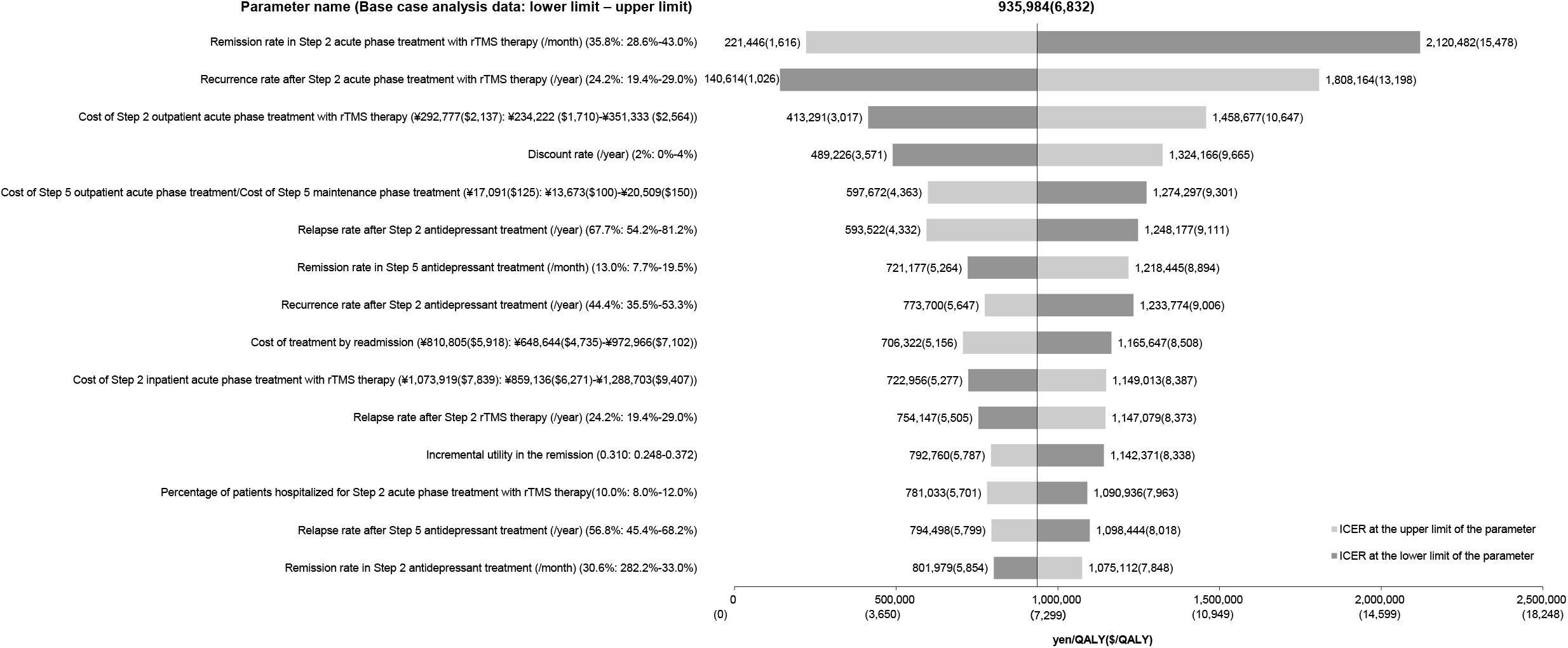
Results of One-way Sensitivity Analysis. The central vertical line indicates the ICER from the base case analysis. The light color bands indicate the results for the parameters changed up to the upper limit of the range. The dark color bands indicate the results for the parameters changed down to the lower limit of the range. Parameters are listed in descending order of the magnitude of impact on the analysis results. Of the parameters set for the model, the remission rate of step 2 acute phase treatment with rTMS therapy was the most impact on the analysis results.

**Figure 3.**
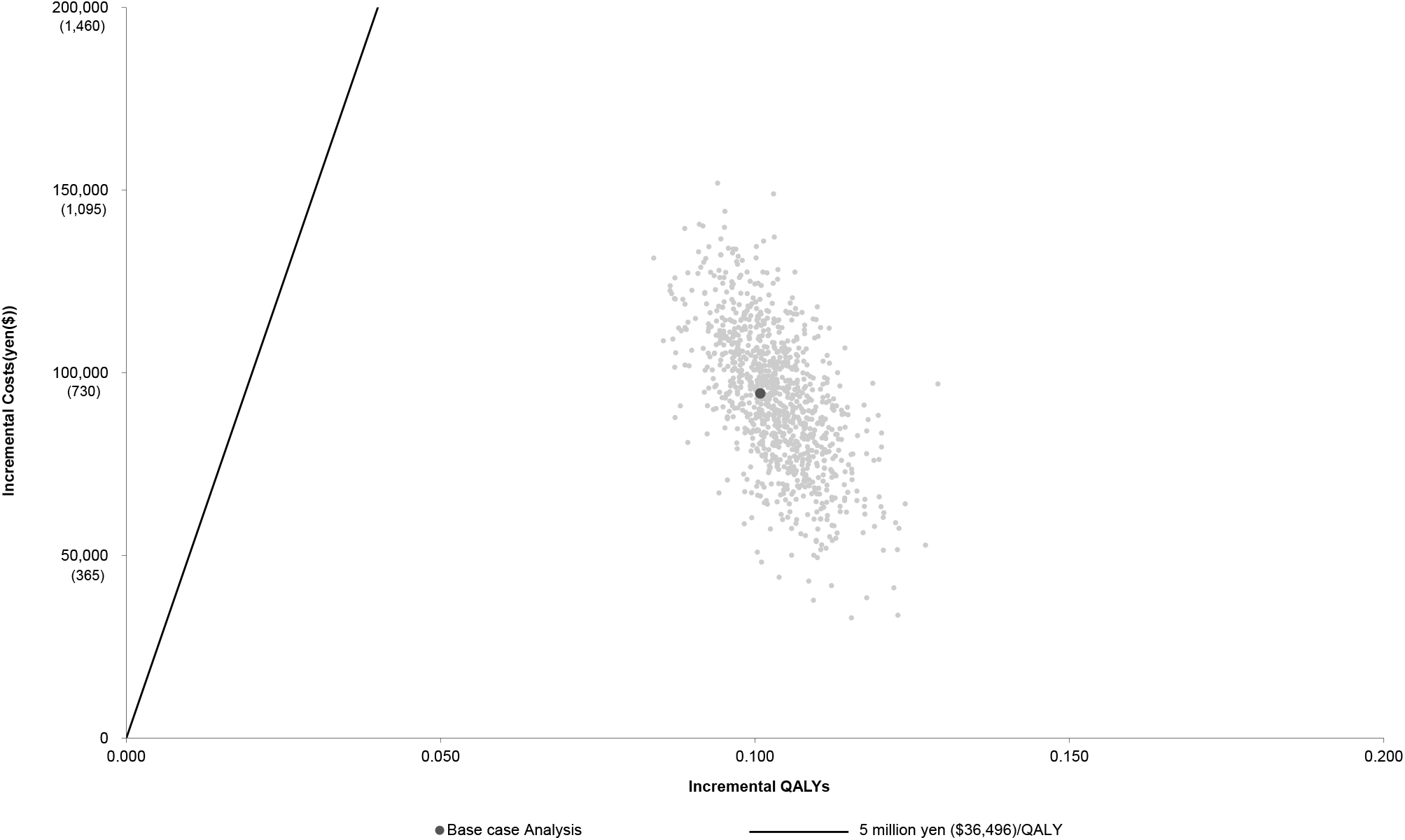
Results of Probabilistic Sensitivity. Analysis The straight line indicates the ICER equal to ¥5 million ($36,496)/QALY. The dark color point indicates the ICER from the base case analysis, and light color points indicate the ICERs when the parameters are varied according to the distribution. The probability of rTMS therapy for TRD to be cost-effective was 100%.

## Discussion

This is the first analytical study in Japan to evaluate the cost-effectiveness of rTMS therapy applied as step 2 acute phase treatment in comparison with antidepressant treatment for patients with TRD managed within the framework of the NHI system in Japan. Since rTMS therapy costs more per visit and requires more frequent visits during the treatment period than antidepressant therapy, the estimated cost during step 2 acute phase treatment was calculated to be ¥382,161 ($2,789) more for rTMS therapy. However, the higher efficacy of rTMS therapy compared with antidepressant treatment, resulting in a higher remission rate with treatment and thus a higher number of patients achieving treatment termination, which in turn reduced the increase in cost for rTMS therapy compared with antidepressant treatment by ¥94,370 ($689). The same result held for effectiveness, with rTMS therapy increasing by 0.101 QALYs because more patients achieved remission during step 2 of the acute phase treatment. Based on these results, the ICER for rTMS therapy compared with antidepressant treatment was calculated to be ¥935,984 ($6,832)/QALY.

The public cost-effectiveness evaluation system in Japan is intended to adjust the reimbursement price under the NHI system. Specifically, under the current public cost-effectiveness evaluation system, reimbursement price reductions are considered when the ICER is ¥5 million ($36,496)/QALY or higher (Center for Outcomes Research and Economic Evaluation for Health, 2019). All the results of each analysis in this study showed that the ICER with rTMS therapy was less than ¥5 million ($36,496)/QALY. Thus, rTMS therapy for patients with TRD was identified to be a cost-effective treatment strategy under the NHI system in Japan.

Since TRD is a disorder that often develops into a chronic condition, the main analysis was conducted as a long-term simulation over the lifetime of patients with TRD; however, a scenario analysis was also conducted with the time horizon changed to 10 years. Although the increase in costs with rTMS compared with antidepressants was greater than that estimated in the base case analysis, the ICER was ¥1,513,265 ($11,046)/QALY, which was substantially lower than ¥5 million ($36,496)/QALY (Table 3). Thus, rTMS therapy was confirmed to be a remarkably cost-effective treatment from a short-term perspective.

In other countries, several reports have been published on the efficacy of maintenance rTMS as a treatment strategy to prevent recurrence in patients who have achieved remission of depressive symptoms with acute phase treatment with rTMS therapy, as well as the efficacy of reintroduction of rTMS therapy at the time of relapse or recurrence (Janicak et al., 2010; Senova et al., 2019). We also evaluated the cost-effectiveness of rTMS, considering the possibility that rTMS could be used as a maintenance phase treatment and reintroduced at the time of relapse or recurrence in the future in Japan. In this context, we conducted the scenario analysis in which acute phase treatment with rTMS is applied again at the time of recurrence for patients after completion of the acute phase of step 2 treatment with rTMS therapy. When applying rTMS therapy 5 times a week to recurrent cases, reintroduction of rTMS therapy was dominant over continued antidepressant treatment since more patients could achieve remission and terminate treatment. In addition, we also conducted the scenario analysis in which maintenance rTMS therapy was applied once a week in the outpatient setting to patients who achieved remission with acute phase of rTMS therapy. The result showed that maintenance rTMS therapy was costly compared with the base case analysis. However, this treatment strategy would still be deemed cost-effective given that the ICER was ¥2,418,868 ($17,656)/QALY, much lower than the reference value of ¥5 million ($36,496)/QALY. Since depression is a disorder susceptible to relapse and recurrence, it is important to manage depression firmly after acute phase treatment as a long-term treatment strategy to prevent refractoriness and chronicity of the condition. Especially, it is urgent to establish effective continuation therapy (i.e., for patients who relapsed after acute phase treatment without achieving remission) and maintenance therapy (i.e., for patients who achieved remission in response to acute phase treatment but require prophylactic therapy to prevent recurrence).

rTMS therapy is an effective treatment with or without concomitant antidepressant treatment; however, it is expected to exert a greater antidepressant effect with concomitant antidepressant treatment than without (Wang et al., 2017). The scenario analysis for patients who have concurrent use of rTMS and antidepressant treatment showed that the strategy was dominant compared with medication alone.

Depression is a disorder that has a significant impact on the productive age population because it is more likely to occur in the working population in middle age (Ministry of Health, Labour and Welfare of Japan, 2020). The decline in labor productivity arising from this clinical-epidemiological characteristic of depression can be directly linked to socioeconomic losses. With this background, we conducted a scenario analysis taking into account the impact of the loss of productivity associated with depression, and the result of the scenario analysis was dominant. Therefore, rTMS therapy can provide a positive impact on society as a whole, in addition to being a viable treatment modality for patients with TRD.

Previous studies evaluating the cost-effectiveness of rTMS therapy in patients with TRD compared with antidepressant treatment include a study by Nguyen et al. in Australia (Nguyen and Gordon, 2015) and Voigt et al. in the United States (Voigt et al., 2017). Since these studies are cost-effectiveness analyses in their respective country’s healthcare environments, they naturally differ from the clinical process of depression under the healthcare environments in Japan. Therefore, in this study, we constructed our own model for treatment steps related to antidepressant therapy and the timing of hospitalization that reflects the healthcare environments for depression treatment in Japan.

While the analysis settings and model structures for cost-effectiveness differed among previous studies, the finding that rTMS therapy is more cost-effective than antidepressant treatment was consistent in all previous studies, and our present analyses are also consistent with those previous findings. On the other hand, the previous studies differed in some respects from our results, showing that rTMS therapy was dominant over antidepressant treatment in terms of higher QALY and lower cost (Nguyen and Gordon, 2015; Voigt et al., 2017). Even though the study by Nguyen et al. had a shorter time horizon of 3 years than the present study, the cost of rTMS therapy was lower than that of antidepressant treatment (Nguyen and Gordon, 2015). Here, potential factors underlying the differences between the results of the previous study and ours may include: (1) providing a sufficient amount of rTMS therapy for more than 6 weeks until remission is achieved during the acute phase of rTMS therapy; (2) providing a booster rTMS therapy for cases that developed recurrence after achieving remission during the acute phase of rTMS therapy; and (3) providing regular rTMS therapy as maintenance phase treatment to prevent recurrence after remission. These approaches may reduce the full-scale worsening of depression, which in turn reduces total costs by preventing subsequent transition to more expensive medical treatment such as hospitalization or ECT. In addition, the following factors may be related to the finding by Voigt et al. that rTMS therapy is more cost-effective compared with antidepressant therapy (Voigt et al., 2017). That is, it is possible that the small cost difference per cycle between rTMS therapy and antidepressant therapy (about $1,000) accentuated the direct impact of the difference in efficacy of the two therapies (rTMS therapy > antidepressant therapy), resulting in an early absorption of the total cost difference between the two therapies.

One-way sensitivity analysis demonstrated that the model was most sensitive to the remission rate of step 2 acute phase treatment with rTMS therapy. In this connection, the previous studies adopted the remission rate of 21.5% reported in the meta-analysis (Nguyen and Gordon, 2015), that is, a lower remission rate than that adopted in step 2 antidepressant therapy in the present study. On the other hand, the remission rate used in the present study was derived from the NeuroStar® registry study (Sackeim et al., 2020). It is possible that the large differences in remission rates to rTMS therapy between studies can be attributed to differences in patient background, number of treatment sessions, stimulation intensity, and clinical assessment methods. However, given that recent rTMS clinical studies have shown higher treatment efficacy compared with the results in the early 2000s, when rTMS therapy was first applied as a new treatment modality mainly in Western countries, improvements in the performance of rTMS devices and treatment protocols in recent studies may have contributed significantly to the increased efficacy (Vogel and Soti, 2022). The NeuroStar® registry study (Sackeim et al., 2020) was a real-world, open-label trial and may have overestimated the remission rate compared with randomized controlled trials. Despite these limitations, the results of this study have a certain degree of validity, as the ICER remained sufficiently lower than ¥5 million ($36,496)/QALY in the one-way sensitivity analysis conducted by varying the remission rate of acute phase rTMS therapy corresponding to that of antidepressant therapy.

As of 2022, a multicenter clinical study is in progress in Japan to examine the effect of maintenance rTMS treatment in preventing recurrence in patients who have responded to acute rTMS treatment, as well as the TMS database registry project to comprehensively collect clinical data related to rTMS therapy in Japanese patients with depression (Noda et al., 2022), which will be important resources for a wide variety of clinical and epidemiological data on the efficacy, safety, and tolerability of rTMS therapy in Japan. Furthermore, it will be possible to utilize such data in the future to conduct cost-effectiveness analyses of rTMS therapy based on real-world data. More widespread application of rTMS therapy as a general treatment option for patients with TRD could lead to optimizing depression treatment strategies and ultimately solving the problem of depression-related productivity loss in the society (Asami et al., 2015; Yamabe et al., 2019). Furthermore, to sustainably provide rTMS therapy, which is a useful therapeutic technique in these diverse aspects, it is crucial to secure and maintain adequate medical resources at healthcare institutions. Currently, however, the reimbursement price for rTMS therapy is set considerably lower in Japan than in other countries (Nguyen and Gordon, 2015; Voigt et al., 2017). In light of this situation, we conducted a threshold analysis of the reimbursement price for rTMS therapy as comparing with the price of other countries. The analysis revealed rTMS was still deemed cost-effective up to ¥30,800 ($225) when ¥5 million/QALY was set as the ICER threshold (Supplementary Figure 2). Thus, if the price for rTMS therapy increased under the NHI, medical institutions in Japan that provide rTMS therapy under the current difficult circumstances can receive compensation that is reasonable enough to facilitate its feasibility, which in turn can contribute to resolving the unmet needs for rTMS therapy in Japan.

The present study has the following limitations: 1) The clinical and utility parameters were extrapolated over the course of the lifetime of a patient based on shorter term data. This method involves some uncertainty when applied to the lifetime cost-effectiveness analysis of rTMS therapy; 2) The clinical parameters were all sourced from overseas study results since there was not sufficient data from large-scale studies conducted in Japan; 3) The utility parameters were all derived from overseas data. They may not faithfully reflect the health state of Japanese patients with depression. Additionally, the methods to estimate utility values were not specified in many previous studies; 4) The cost for rTMS therapy was estimated based on the expert opinions since there was not sufficient data for rTMS on the Japanese Claims Database. The rTMS therapy had started to be reimbursed by the NHI recently; 5) Some parameters were set based on the expert opinions of the authors in the field. Thus, these parameters may not fully reflect the clinical practice of depression in Japan.

## Conclusion

The present study suggested that rTMS therapy for TRD would be cost-effective compared with antidepressant therapy. If rTMS therapy can be provided to more patients with TRD in a more appropriate form, it will lead to further streamlining and optimization of treatment strategies for TRD in medical care in Japan.

## Supporting information

checklist

## Data Availability

All data produced are available online

## Contributors

All authors gave substantial contribution to this work. YN and CM conceived the concept of this research. YN, CM, and YK drafted the manuscript. YN, SK, and MM provided interpretation of this research and revised the manuscript. All authors have reviewed and approved the manuscript. The corresponding author had full access to all the data in the study and had final responsibility for the decision to submit for publication.

## Acknowledgement

We would like to acknowledge Keigo Hanada and Yuta Fukuoka (Crecon Medical Assessment, Tokyo, Japan) for their assistance in the professional analyses of this study. YN has received a Grant-in-Aid for Scientific Research (B) (21H02813) from the Japan Society for the Promotion of Science (JSPS), research grants from Japan Agency for Medical Research and Development (AMED), investigator-initiated clinical study grants from Teijin Pharma Ltd, and Inter Reha Co., Ltd. He has also received research grants from Daiichi Sankyo Scholarship Donation Program. He has received speaker’s honoraria from Sumitomo Pharma Co., Ltd, Qol Co., Ltd, Teijin Pharma Ltd., and Takeda Pharmaceutical Co., Ltd. He also receives equipment-in-kind support for an investigator-initiated study from Magventure Inc., Inter Reha Co., Ltd., and Miyuki Giken Co., Ltd. SK has received research grants and/or speaker’s honoraria from Inter Reha Co., Ltd., Kyowa Pharmaceutical Industry Co., Ltd., Lundbeck Japan K.K., Sumitomo Pharma Co., Ltd., Otsuka Pharmaceutical Co., Ltd., Takeda Pharmaceutical Co., Ltd., Teijin Pharma Ltd., and Viatris Inc. MM has received grants and/or speaker’s honoraria from Asahi Kasei Pharma, Astellas Pharma, Daiichi Sankyo, Sumitomo Pharma, Eisai, Eli Lilly, Fuji Film RI Pharma, Janssen Pharmaceutical, Kracie, Meiji-Seika Pharma, Mochida Pharmaceutical, Merck Sharp and Dohme, Novartis Pharma, Ono Pharmaceutical, Otsuka Pharmaceutical, Pfizer, Shionogi, Takeda Pharmaceutical, Teijin Pharma Ltd., Mitsubishi Tanabe Pharma, and Yoshitomi Yakuhin.

