## Supplementary for "Cost-effectiveness analysis comparing repetitive transcranial magnetic stimulation therapy with antidepressant treatment in patients with treatment-resistant depression in Japan"

### Figure Legends

**Supplementary Figure 1. Patient flow in cost analysis using the JMDC Claims Database**

The medical cost for TRD patients was calculated with the use of the JMDC Claims Database provided by JMDC Inc. Medical cost was calculated on the 4,211 TRD patients selected for this analysis using the inclusion/exclusion criteria from the 245,871 patients diagnosed as having depression between July 2016 and June 2019.

**Supplementary Figure 2. Results of analysis at varying amounts reimbursed for each session of rTMS therapy**

ICER was ¥1,428,627 ($10,428)/QALY when the price for rTMS therapy used in the study by Nguyen et al. (150AUD: ¥14,286) was adopted into this model. ICER was ¥4,650,374 ($33,944)/QALY when the prices for rTMS therapy used in the study by Voigt et al. (first session $367: ¥50,279, second and subsequent sessions $206: ¥28,222) were adopted. Reimbursement price for one session of rTMS therapy was identified to be ¥30,800 ($225) at the ICER threshold of ¥5 million/QALY.

### Supplementary Figure 1. Patient flow in cost analysis using the JMDC Claims Database

Patients for whom care records during the study period are available **245,871**

| Patients not satifying the inclusion criteria ① | **206,789** |
|---|---|
| - No definite diagnosis of depression during the study period | **5,941** |
| - Three or more drugs used for the primary treatment | **85,739** |
| - No secondary treatment received within 30 days after the estimated last drug dose date during the primary treatment | **206,789** |

Patients with the index date **39,082**

| Patients not satisfying the inclusion criteria ② | **11,227** |
|---|---|
| - No psychotropic prescription or admission to psychiatric ward during the follow-up period | **10,208** |
| - No psychotropic prescription during the follow-up period | **10,336** |
| - No admission to the psychiatric ward during the follow-up period | **36,844** |
| - Age less than 18 years at the time of index date | **1,204** |

Patients satisfying the inclusion criteria **27,855**

| Patients falling under any of the exclusion criteria | **23,644** |
|---|---|
| - Definitely diagnosed as having severe depressive episode, mania, schizophrenia, epilepsy, convulsive attack, intracranial lesion, etc. accompanied by psychotic symptoms before the index date | **16,947** |
| - Definitely diagnosed as being pregnant during the follow-up period | **205** |
| - Having received medical procedure with implant type medical device, etc. contraindicated for rTMS during the follow-up perio | **403** |
| - The study period does not cover the 180 days before the earliest definite diagnosis of depression | **18,359** |
| - The baseline period cannot be secured | **9,534** |
| - Follow-up period is less than 1 month | **0** |
| - Records of admission available at the time of index date | **275** |

Patients falling under none of the exclusion criteria **4,211**

Patients included in the analysis **4,211**

### Supplementary Figure 2. Results of analysis at varying amounts reimbursed for each session of rTMS therapy

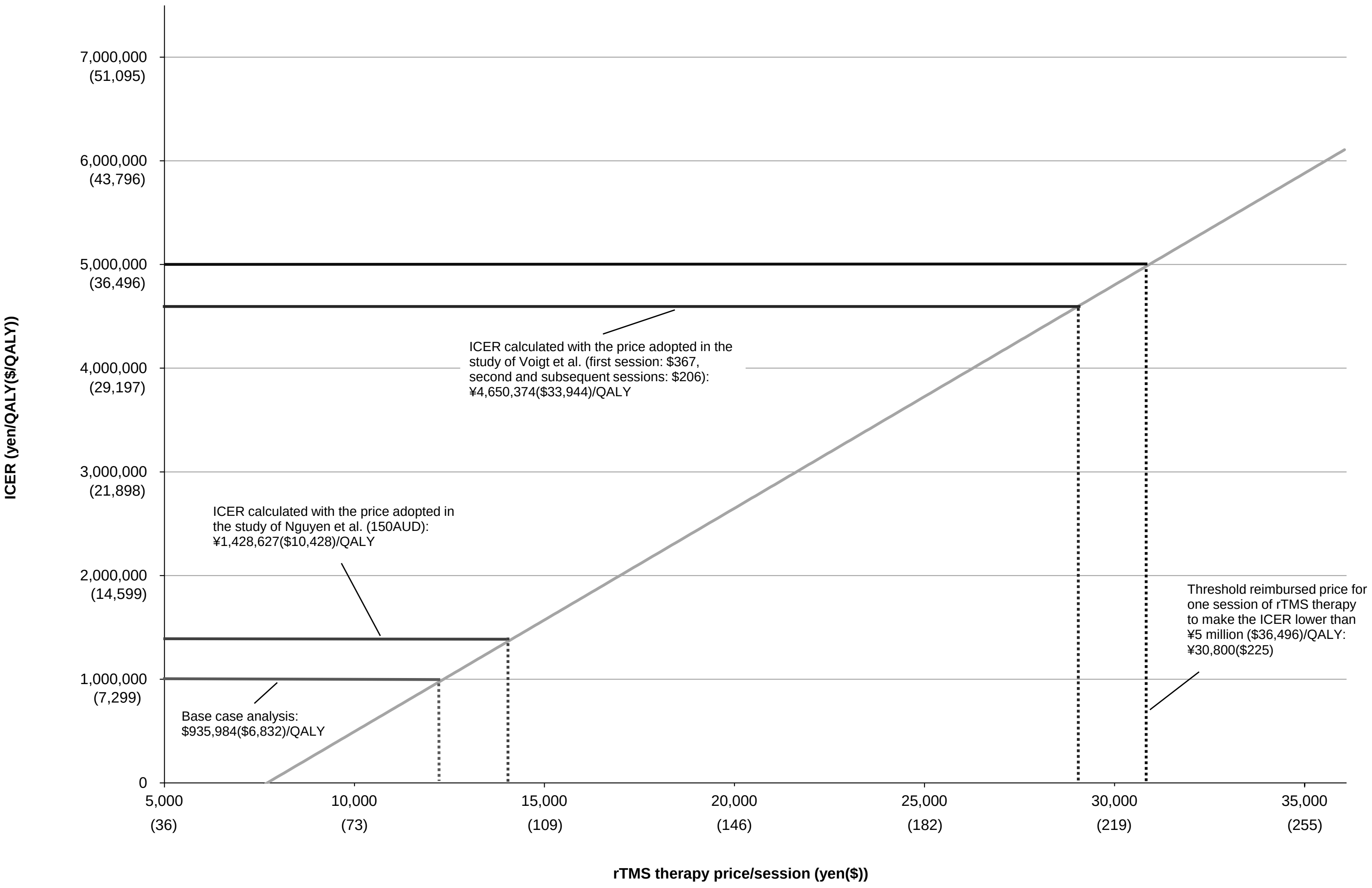

**Supplementary Table 1. Summary of cost analysis using the JMDC Claims Database**

| Variable | Definition |
|---|---|
| Study period | July 2016 to June 2019 |
| Index date | Secondary treatment starting date (date of antidepressant prescription within 30 days after the end of first treatment) |
| Follow-up period | From the index date for each patient to the last day with the record on the JMDC database |
| Duration of treatment | From the date of estimated last intake of antidepressant or the drug for augmentation therapy (date of prescription + number of days covered by prescription) to a point of time by which 31 days or more were without identifiable prescription of these drugs |
| Patients studied | |
| Inclusion | Diagnosed as having depression during the study period (Supplementary Table 2) and having begun to receive antidepressant treatment defined in Supplementary Table 3 after the diagnosis |
| | Having received secondary treatment within 30 days after the date of the last prescription for the primary treatment + the number of days covered by prescription (estimated date of last drug intake) |
| | Having received prescription of any of the psychotropics defined in Supplementary Table 3 or being admitted to a psychiatric ward during the follow-up period |
| Exclusion | Having been diagnosed as having severe depressive episode, mania, schizophrenia, epilepsy, convulsive attack, intracranial lesion, etc. accompanied by psychotic symptoms before the index date |
| | Having received medical procedure using a metallic or other device or any other material applied to the head or within 30 cm from the head during the follow-up period |
| | Having been diagnosed as being pregnant during the follow-up period |
| Data collected | The data on the following costs were collected during the treatment continued period |
| Drug costs at each treatment step | Calculated by averaging (weighted by the percentage of each drug prescribed) of the daily prescribed quantity of antidepressant or drug for augmentation therapy at each treatment line x drug price x monthly frequency of dosing |
| Inpatient care costs | Monthly costs of inpatient care after admission for a reason of depression |
| Outpatient care costs | Costs of outpatient care accompanied by prescription of antidepressant or drug for augmentation therapy |
| Outpatient test costs | Costs of tests conducted during outpatient visit accompanied by prescription of antidepressant or drug for augmentation therapy |
| ECT costs | Monthly costs of ECT applied to inpatients (192 cases) admitted after diagnosis of depression |

### Supplementary Table 2. Definition of depression

| Variable | Definition | |
|---|---|---|
| | **ICD10 code** | **ICD10 name** |
| Moderate or severer depressive episode | F321 | Moderate depressive episode |
| | F322 | Severe depressive episode without psychotic symptoms |
| Moderate or severer recurrent depressive disorder | F331 | Recurrent depressive disorder, current episode moderate |
| | F332 | Recurrent depressive disorder, current severe episode without psychotic symptoms |
| Depressive episode, unspecified | F329 | Depressive episode, unspecified |
| Recurrent depressive disorder, unspecified | F339 | Recurrent depressive disorder, unspecified |

### Supplementary Table 3. Definition of antidepressants/psychotropics

| Classification | Active Ingredient |
|---|---|
| Antidepressants | |
| Tricyclic antidepressants | Amitriptyline hydrochloride |
| | Amoxapine |
| | Imipramine hydrochloride |
| | Clomipramine hydrochloride |
| | Dosulepin hydrochloride |
| | Trimipramine maleate |
| | Nortriptyline hydrochloride |
| | Lofepramine hydrochloride |
| Tetracyclic antidepressants | Setiptiline maleate |
| | Maprotiline hydrochloride |
| | Mianserin hydrochloride |
| SSRI | Escitalopram oxalate |
| | Sertraline hydrochloride |
| | Paroxetine hydrochloride |
| | Fluvoxamine maleate |
| SNRI | Duloxetine hydrochloride |
| | Milnacipran hydrochloride |
| | Venlafaxine hydrochloride |
| SARI | Trazodone hydrochloride |
| Serotonin reuptake inhibitors/serotonin receptor modulators | Vortioxetine hydrobromide |
| NaSSA | Mirtazapine |
| Psychotropics | |
| Tricyclic antidepressants | Amitriptyline hydrochloride |
| | Amoxapine |
| | Imipramine hydrochloride |
| | Clomipramine hydrochloride |
| | Dosulepin hydrochloride |
| | Trimipramine maleate |
| | Nortriptyline hydrochloride |
| | Lofepramine hydrochloride |
| Tetracyclic antidepressants | Setiptiline maleate |
| | Maprotiline hydrochloride |
| | Mianserin hydrochloride |
| SARI | Trazodone hydrochloride |
| Lithium | Lithium carbonate |
| T3/T4 preparations | Levothyroxine sodium |
| | Dry thyroid |
| | Liothyronine sodium |
| Aripiprazole | Aripiprazole |
| Quetiapine | Quetiapine fumarate |
| Olanzapine | Olanzapine |
| Risperidone | Risperidone |

**Supplementary Table 4. List of parameters used for scenario analysis**

| Parameter | Scenario analysis value | Source |
|---|---|---|
| 1) rTMS therapy continued as Step 2 maintenance phase treatment | | |
| Relapse rate in Step 2 maintenance phase treatment with rTMS therapy | 9.1%/year | (Haesebaert et al., 2018) |
| Recurrence rate in Step 2 maintenance phase treatment with rTMS therapy | 24.2%/year | (Wang et al., 2017) |
| Cost of Step 2 maintenance phase treatment with rTMS therapy | ¥69,379 ($506)/month | JMDC Claims Database, Expert opinion |
| 2) rTMS therapy applied as Step 3 acute phase treatment upon recurrence following Step 2 rTMS therapy | | |
| Response rate in Step 3 acute phase treatment with rTMS therapy | 84.2%/month | (Janicak et al., 2010) |
| Remission rate in Step 3 acute phase treatment with rTMS therapy | 35.8%/month | (Sackeim et al., 2020) |
| Relapse rate after to Step 3 acute phase treatment with rTMS therapy | 24.2%/year | (Wang et al., 2017) |
| Recurrence rate after Step 3 acute phase treatment with rTMS therapy | 24.2%/year | (Wang et al., 2017) |
| Cost of Step 3 acute phase treatment with rTMS therapy | ¥370,892 ($2,707)/month | JMDC Claims Database, Expert opinion |
| 3) rTMS therapy plus antidepressant treatment applied as Step 2 acute phase treatment | | |
| Response rate in Step 2 acute phase treatment with rTMS plus antidepressant treatment | 68.9%/month | (Sackeim et al., 2020) |
| Remission rate in Step 2 acute phase treatment with rTMS plus antidepressant treatment | 35.8%/month | (Sackeim et al., 2020) |
| Relapse rate after Step 2 acute phase treatment with rTMS plus antidepressant treatment | 15.9%/year | (Wang et al., 2017) |
| Recurrence rate after Step 2 acute phase treatment with rTMS plus antidepressant treatment | 15.9%/year | (Wang et al., 2017) |
| Cost of Step 2 acute phase treatment with rTMS plus antidepressant treatment | ¥384,975 ($2,810)/month | JMDC Claims Database, Expert opinion |
| 4) Productivity loss due to depression taken into consideration | | |
| Employment rate among patients with depression | 60.40% | (Statistics Bureau of Japan, 2021) |
| During outpatient treatment | | |
| Absenteeism | 14.10% | (Yamabe et al., 2019) |
| Presenteeism | 42.70% | (Yamabe et al., 2019) |
| During inpatient treatment | | |
| Absenteeism | 100% | Assumption |
| Presenteeism | 100% | Assumption |
| Mean wage (annual) | | |
| 40-44 years | ¥5,323,700 ($38,859) | (Ministry of Health, Labour and Welfare of Japan, 2021b) |
| 45-49 years | ¥5,552,900 ($40,532) | |
| 50-54 years | ¥5,887,100 ($42,972) | |
| 55-59 years | ¥5,807,500 ($42,391) | |
| 60-64 years | ¥4,356,100 ($31,796) | |
| 65-69 years | ¥3,615,700 ($26,392) | |
| 70 years or older | ¥3,264,100 ($23,826) | |
